# How often is the core outcome set for low back pain used in clinical trials? A protocol for a meta-epidemiological study

**DOI:** 10.1101/2023.01.11.23284425

**Authors:** Tiziano Innocenti, Stefano Salvioli, Patricia Logullo, Silvia Giagio, Raymond Ostelo, Alessandro Chiarotto

## Abstract

**Background:** Non-specific Low back pain (NSLBP) is the worldwide leading cause of disability, accounting for large costs for healthcare systems and work productivity. Many treatment options are available for patients with NSLBP. Authors of systematic reviews on LBP report that outcomes are often measured and reported inconsistently. This inconsistency limits the comparison of findings among trials, and it can be due to selective outcome reporting bias (e.g. reporting only outcomes with positive results in a publication), which strongly affects the conclusions of systematic reviews. Recommendations for standardised reporting of outcome measurement instruments in clinical studies were initially publicated in 1998 and updated through an international consensus Delphi study by Chiarotto and colleagues in 2015. This updated Core Outcome Set (COS) for NSLBP included the following core outcome domains: “physical functioning”, “pain intensity”, “health-related quality of life”, and “number of deaths”. With the exception of “number of deaths”, the other three core domains were already included in the core set publicated in 1998 by Deyo et al. In 2018, another international consensus of Chiarotto et al. formulated recommendations on which core outcome measurement instruments (Core Outcome Measurement Set – COMS) should be used in NSLBP trials. A consensus was reached on Numeric Rating Scale (NRS) for “pain intensity”, Oswestry Disability Index (ODI) or Roland-Morris Disability Questionnaire (RMDQ-24) for “physical functioning”, Short Form Health Survey 12 (SF12) or 10-item PROMIS Global Health (PROMIS-GH-10) for “HRQOL”. Therefore, the recommended COS has been in the public domain for more than 20 years. However, it is still unknown whether it has changed the selection of outcomes used in NSLBP trials during this period.

**Objectives:** (1)To assess the uptake of the COS for NSLBP in clinical trials; (2)To assess the uptake of the Core Outcome Measurement Set for NSLBP in clinical trials; (3)To analyse whether specific study characteristics (year of registration, sample size, country of origin, duration of follow-up, phase of the trial, intervention, and source of funding) are associated with the COS uptake

**Methods:** We will adopt Kirkham et al.’s recommendations on the assessment of COS uptake. We will search the World Health Organization (WHO) International Clinical Trials Registry Platform (ICTRP) and Clinicaltrials.gov registry to identify potentially eligible trial protocols. Two reviewers (TI and SG) will select potentially eligible entries and evaluate whether they meet the eligibility criteria. A consensus meeting will be held to determine agreement on the selection; in case of disagreement, a third reviewer (SS) will decide on inclusion. We will calculate the percentage of clinical trials that planned to measure data on the NSLBP full COS. We will also calculate the proportion of trials that reported the percentage of trials measuring the full COS per year. We will calculate the percentage of the NSLBP core outcome measurement instruments used per each domain described in the COS. Lastly, we will perform a multivariable logistic regression analysis to assess the relationship between the full COS uptake (yes/no) as the dependent variable and the following independent variables: year of registration, sample size, country of origin, duration of follow-up interval, phase of the trial (III or IV), intervention (pharmacological trial vs non-pharmacological trial), and source of funding (commercial vs non-commercial vs no funding).

**Ethics and dissemination:** A manuscript will be prepared and submitted for publication in an appropriate peer-reviewed journal upon study completion. We believe that the results of this investigation will be relevant to researchers paying more attention to the synthesis of the evidence to translate clinical implications to key stakeholders (healthcare providers and patients).

## INTRODUCTION

Low back pain (LBP) is the worldwide leading cause of disability, accounting for high costs for healthcare systems and work productivity^1,2^. The large majority of patients with LBP are labelled as having non-specific LBP (NSLBP) that is defined as “tension, soreness and/or stiffness in the lower. back region for which it is not possible to identify a specific cause of the pain” ^3,4^.

Many treatment options are available for patients with NSLBP^5^. Randomised controlled trial (RCT) are the best study designs to provide accurate estimates of the effectiveness of an intervention by comparing its relative effects on outcomes chosen to identify benefits or harms^6^. It is essential that outcomes reported in trials are those considered meaningful by all stakeholders (i.e. patients, clinicians, and all those involved in the process of care) consider meaningful outcomes ^7^. However, authors of systematic reviews on LBP report that outcomes are often measured and reported inconsistently across trials^8,9^. This inconsistency limits the comparison of findings between trials^10^, and it can be due to selective outcome reporting bias (e.g. reporting only outcomes with positive results in a publication), which strongly affects conclusions of systematic reviews^11^.

These issues could be addressed through the development and use of an agreed and standardized set of outcomes^10^, known as a core outcome set (COS), which should be measured and reported in all trials for a certain health condition^12^. In the field of LBP, recommendations for standardized reporting of outcomes in clinical studies were published in 1998^13^ and updated through an international consensus Delphi study led by Chiarotto and colleagues in 2015^14^. This updated COS for NSLBP included the following core outcome domains: “physical functioning”, “pain intensity”, “health-related quality of life” (HRQOL), and “number of deaths”^14^.

Besides defining what outcomes should be investigated, it is necessary to agree on the best tools to measure them, so that trials can use them in a standardised and comparable way. For this purpose, in 2018, another international consensus led by Chiarotto et al. formulated recommendations on which core outcome measurement instruments (Core Outcome Measurement Set – COMS) should be used in NSLBP trials^15^. A consensus was reached on the Numeric Rating Scale (NRS) for “pain intensity”, Oswestry Disability Index (ODI) or Roland-Morris Disability Questionnaire (RMDQ-24) for “physical functioning”, Short Form Health Survey 12 (SF12) or 10-item PROMIS Global Health (PROMIS-GH-10) for “HRQOL”, and a simple statement on the number of deaths occurring in the trial for “number of deaths”. With the exception of “number of deaths”, the other three core domains (and measurement instruments) were already included in the core set published in 1998 by Deyo et al.^13^ Therefore, for the purposes of this study, we will consider the domains and the instruments that are included both in the Deyo’s and Chiarotto’s COS.

The recommended COS has been in the public domain for more than 20 years^13^, but it is still unknown whether authors have changed the selection of outcomes used in NSLBP trials during this period.

Therefore, the objectives of our meta-epidemiological study are the following:

### Primary Objective

- To assess the uptake of the COS in NSLBP clinical trials;

### Secondary Objectives

- To assess the uptake of the COMS for NSLBP;
- To analyse whether specific study characteristics (year of registration, sample size, country of origin, duration of follow-up, phase of the trial, intervention, and source of funding) are associated with the COS uptake.

## MATERIAL AND METHODS

We will follow and adapt the PRISMA 2020 checklist^16^ for the reporting of this manuscript (or a specific reporting checklist for meta-epidemiological studies, if available at the time of reporting^17^). We will adopt Kirkham et al.’s recommendations on the assessment of COS uptake^18^ for our primary analysis.

### Data source and search strategies

We will search for trials registry entry on NSLPB the World Health Organization (WHO) International Clinical Trials Registry Platform (ICTRP) through its research portal (https://trialsearch.who.int) and Clinicaltrials.gov registry. The search was performed on November 30^th^, 2022 without time/language restrictions. The Clinical Trials Search Portal provides access to a WHO central database containing the trial registration data sets provided by the registries listed in table 1. The search will be conducted using the “advance search” options as the following:

**Table 1:**
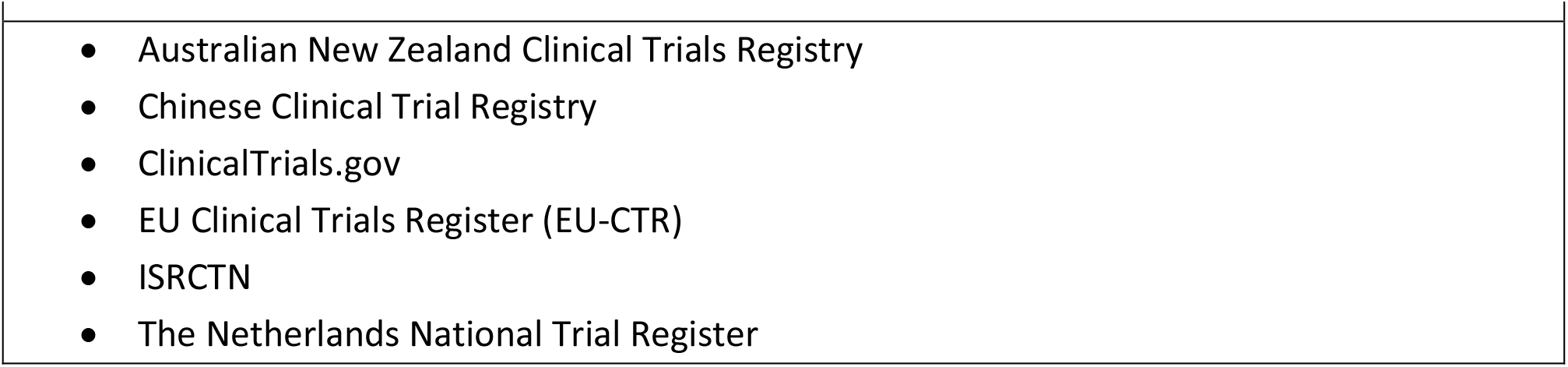
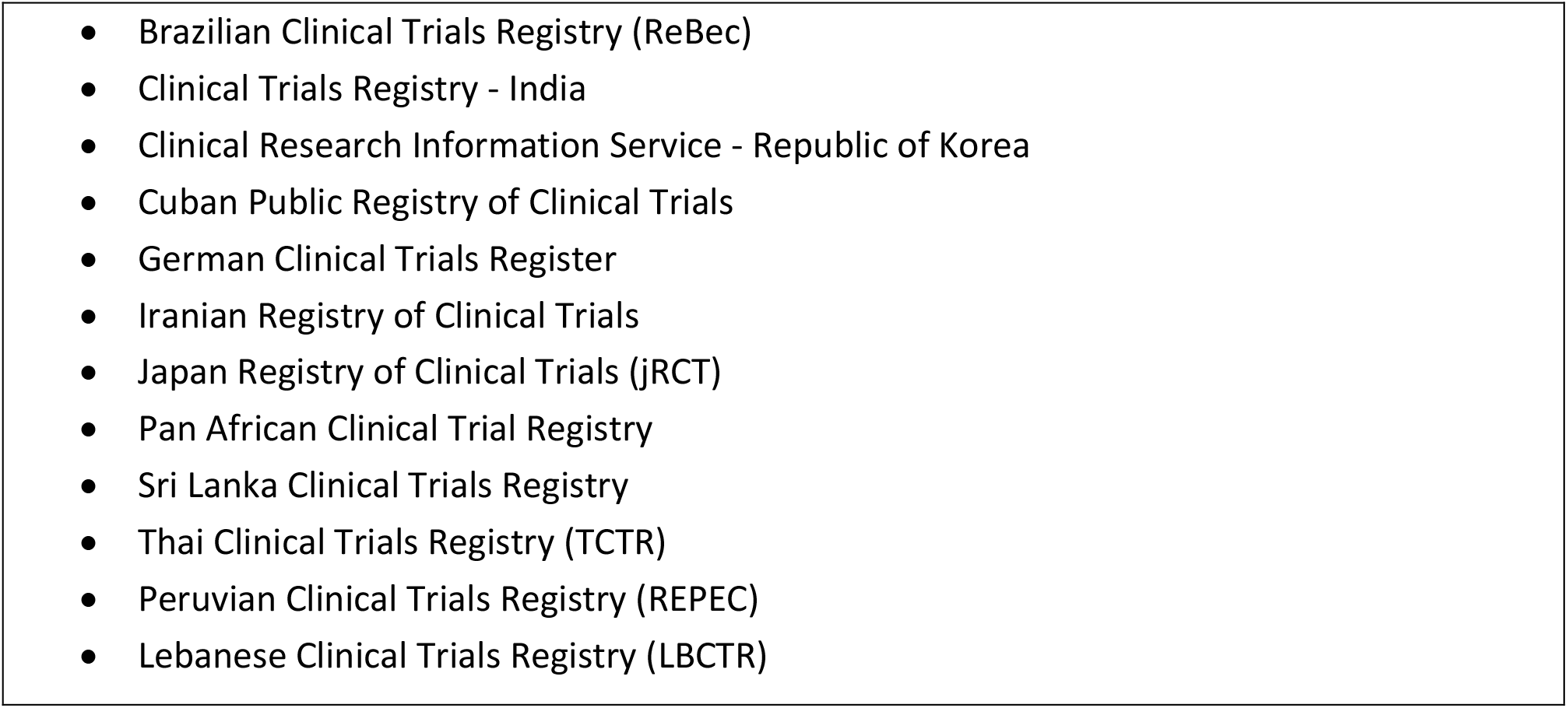
Registries included in the World Health Organization International Clinical Trials Registry Platform

- “Low back pain” in the title
- Recruitment status is “ALL”
- Phases are: “Phase 3 and Phase 4”

To identify potentially relevant trials, we will apply the following filters on Clinicaltrials.gov: “conditions: low back pain”, “study type: interventional studies”, and “phase: 3 and 4”.

### Eligibility criteria

The following inclusion criteria will be applied to retrieve eligible trial registry entries: (1) conducted in adult patients (>18 years old) with NSLBP, (2) RCT design assessing effectiveness or efficacy of interventions as defined by Deyo^13^ and Chiarotto et al.^14^

The following exclusion criteria will be used: (1) trials assessing medication dosage as main outcomes or studies focused on safety, not effectiveness or efficacy, (2) RCTs including a “mixed” population of patients with NSLBP and other musculoskeletal disorders (e.g., specific LBP, neck pain); these will be included only if at least 75% of the patients displayed NSLBP.

### Registry entries selection

Registry entries will be exported from ICTRP in XML format. Then, two reviewers (TI and SG) will select potentially eligible entries and evaluate whether they meet the eligibility criteria. A consensus meeting will be held to determine agreement on the selection; in case of disagreement, a third reviewer (SS) will decide on inclusion. The study selection process will be summarized through a flowchart. Rayyan QCRI systematic review software^19^ will be used to carry out the selection process.

### Data extraction

For each eligible trial registry entry, two reviewers independently (TI and SS) will extract information on all planned trial outcomes and assess whether the full NSLBP core outcome set^14^ is listed together with the instruments used. We will consider “full COS” the following outcome domains: “physical functioning”, “pain intensity”, and “health-related quality of life”. We will not include the outcome “death” in this study, since it is a rare event in the prognosis of NSLBP^20,21^ and also the 2015 COS Steering Committee acknowledges that “a short statement, such as ‘‘no deaths occurred in this clinical trial’’, would suffice to cover this outcome domain”^14^.

If trialists had registered a composite outcome, we will consider all the individual outcomes in the composite in the assessment, even if they will not be listed separately, following Kirkham’s approach^18^. We will also collect the following data: (1) year of trial registration; (2) country of origin; (3) planned sample size; (4) duration of follow-up; (5) the intervention type under investigation (pharmacological or non-pharmacological trial); (6) phase of the trial; (7) recruitment status; (8) source of funding (commercial or non-commercial or no funding); (9) trial publication status To check the trial publication status (i.e. whether the protocol retrieved has results published), we will search the Medline and Embase databases using protocol title, authors and Registration ID as search terms. A third reviewer (AC), who was involved in the development of COS^14^ will solve any discrepancy.

#### Data analysis

Descriptive data of the included studies will be reported in tables as mean (standard deviations) or median (interquartile range) frequencies. The following analyses will be performed:

#### Primary analysis

- We will calculate the percentage of studies that planned to measure the full NSLBP COS out of the total sample of trials registry entry included. We will also calculate the percentage of trials that reported the full domain set per year. These will be assessed without time restriction from registries inception to November 30^th^ 2022 to determine change over time. This range is chosen to investigate the trend in the use of COS, starting from a period of time without any COS published (from inception to Deyo 1998^13^) and investigating if Deyo 1998^13^ and Chiarotto 2015^14^ have changed the trend. Graphs will be used to display the change over time.

#### Secondary analysis

- We will report which is the most commonly reported COS domain among the protocols that do not plan to measure the full NSLPB COS (i.e. the protocols in which only one or two outcome domains are reported).
- We will calculate the percentage of the NSLBP COMS^15^ used per each domain described in the COS.
- We will perform a multivariable logistic regression analysis to assess the relationship between the full COS domain uptake (yes/no) as the dependent variable and the following independent variables: year of registration, sample size, country of origin, duration of followup interval, phase of the trial (III or IV), intervention (pharmacological trial vs non-pharmacological trial), and source of funding (commercial vs non-commercial vs no funding). Data will be presented as OR with 95% Confidence interval. A 2-sided p-value<0.05 will be deemed as indicating statistical significance. The assumptions of linearity, homoscedasticity, independence and normality will be checked.

Analyses will be performed using SPSS software (IBM SPSS Statistics for Macintosh, Version 28.0. Armonk, NY: IBM Corp).

## Data Availability

All data produced in the present work will be contained in the manuscript

## ETHICS AND DISSEMINATION

This study does not require an ethics review as we will not collect personal data; it will summarise information from publicly available studies.

A manuscript will be prepared and submitted for publication in an appropriate peer-reviewed journal upon study completion. These meta-epidemiological study findings will be disseminated at a relevant general scientific conference or scientific events in musculoskeletal rehabilitation and research methods. We believe that the results of this investigation will be relevant to researchers interested in evidence syntheses to translate clinical implications to key stakeholders (healthcare providers and patients).

## AUTHOR CONTRIBUTIONS

TI, AC, and RO conceived and designed the study protocol. TI, SS, SG, RO and AC were involved in conceptualising the study objectives and providing input into study selection criteria and plans for data extraction. PL revised and edited the study protocol manuscript. All the authors approved the final version of the protocol.

